# ACHD Care Wheel: Stakeholder- and Theory-Driven Framework to Improve Specialized Adult Congenital Heart Disease Care

**DOI:** 10.1101/2025.04.01.25325065

**Authors:** Anushree Agarwal, Katelyn Macholl, Sedona L. Koenders, Alice Qian, Juhi Mehta, Pranav Ahuja, Karina Buenrostro, Kevin Sun, Daniel Dohan, Michelle Gurvitz, Megumi J. Okumura

## Abstract

**Background:** Patients with adult congenital heart disease (ACHD) experience significant gaps in guideline-recommended specialized care, leading to higher morbidity and mortality. We evaluated barriers and facilitators to specialized care, with particular focus on understanding how to design and evaluate holistic, patient-centered solutions.

**Methods:** We purposely sampled demographically and geographically diverse group of ACHD patients and clinicians in the United States. We conducted semi-structured interviews guided by the Capability, Opportunity, Motivation, and Behavior (COM-B) model and Theoretical Domains Framework (TDF) and analyzed using rapid qualitative analysis.

**Results:** We interviewed 54 participants (37 patients and 17 clinicians). The patient median age was 32 years; 57% were women, 62% were people of color, and 35% had gaps in care of ≥3 years. Findings were organized into six interdependent categories: healthcare system and institutional structure; knowledge and education; sources of support; personal development; identity and personal resources; and ACHD’s placement in one’s life. Two novel findings emerged: a paradoxical facilitating role of parental language barriers and the complex impact of caregiver support on patients’ independent care skills. We developed the ACHD Care Wheel, a theory-guided framework to support the design and evaluation of future interventions.

**Conclusion:** The ACHD Care Wheel is a stakeholder-derived, theory-grounded framework to develop and evaluate interventions to improve specialized ACHD care. Interventions targeting the three priority categories (healthcare system and institutional structure, knowledge and education, and sources of support) are likely to produce effects across multiple domains simultaneously, offering an efficient and theoretically grounded path forward for this growing population.

## I. Introduction

Improvements in pediatric congenital heart care have led to a rapid rise in the number of adults with congenital heart disease (CHD), making this an increasingly urgent public health issue.^1^ It is estimated that over 2 million people with adult congenital heart disease (ACHD) currently live in the United States, with prevalence increasing by 40,000–50,000 every year.^2^ Despite guidelines recommending that patients with ACHD establish and maintain lifelong care with ACHD specialists^3^, up to 85% experience gaps in receiving specialized care.^4–6^ These gaps in care are associated with higher rates of morbidity and mortality, poor quality of life, and a larger burden on an already resource-constrained healthcare system.^7–9^ Interventions to address these gaps are needed, yet there remains a limited understanding of what interventions are effective and how they should be designed and evaluated.

Prior studies using survey-based, administrative, or claims data have identified patient, clinician, and healthcare system barriers to specialized ACHD care, and some have proposed and tested solutions to overcome them.^10–12^ However, these studies lack the in-depth perspective needed to understand why and how barriers affect care engagement, or what solutions could realistically address them. Even when solutions have been proposed, such as a dedicated transition program, there remains significant variability in their adoption in practice,^13^ and it is not well understood how or why any given solution is or is not effective. This limits the ability to design interventions that are scalable, sustainable, and rigorously evaluable. Behavioral theoretical frameworks are increasingly used to clarify the determinants of health behavior and guide intervention design across chronic conditions, yet their application in ACHD remains largely unexplored.^14–16^

To address these gaps, we conducted semi-structured interviews with a geographically and demographically diverse group of ACHD patients and clinicians to identify barriers, facilitators, and solutions for improving specialized ACHD care. We then describe a novel, stakeholder-derived and theory-grounded framework that clarifies the behavioral mechanisms through which these solutions could enhance specialized care and guide the design and evaluation of future interventions.

## II. Methods

### Theoretical Framework

Implementation Science frameworks have been designed to provide a structured approach for planning, implementing, and evaluating interventions to improve health outcomes. The Capability, Opportunity, Motivation–Behavior (COM-B) model and the Theoretical Domains Framework (TDF)^17^ -which target patient, provider and system barriers-have been used to effectively improve guideline-adherent care among patients with cardiac and noncardiac chronic conditions (**Figure 1**).^18–21^ However, these frameworks remain under-utilized in the ACHD population.

**Figure 1:**
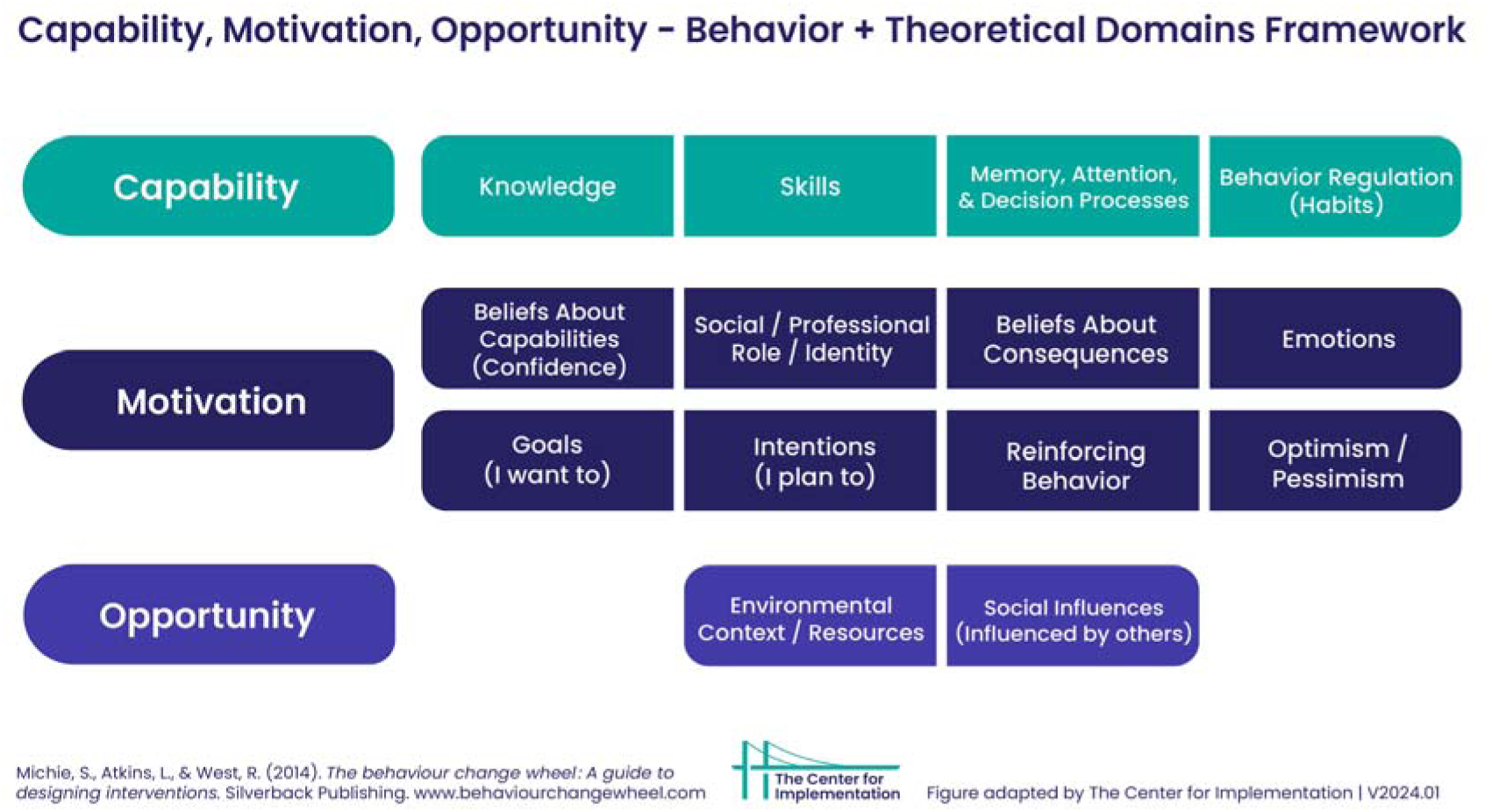
Capability, Opportunity, Motivation - Behavior & Theoretical Domains Framework. Adapted from Michie, S., van Stralen, M. M., & West, R. (2011). The behaviour change wheel: A new method for characterising and designing behaviour change interventions. Implementation Science, 6, 42. https://doi.org/10.1186/1748-5908-6-42. Image adapted by The Center for Implementation, © 2023. Version: V2024.01. https://thecenterforimplementation.com/toolbox/com-b-tdf

The COM-B model outlines the roles and importance of capability, motivation, and opportunity to change specific behaviors.^20^ TDF further elaborates on the COM-B model by identifying 14 sources of behavior, describing different types of individual barriers and facilitators, and tying each back to one of the three COM-B model constructs.^18^ We utilized patient activation and engagement, a concept rooted in the chronic care model, as the target behavior to improve specialized lifelong ACHD care, given its emphasis on supporting patients as active partners with healthcare systems.^22^ Higher patient activation has been associated with greater adherence to chronic care behaviors, reduced hospitalizations and emergency department utilization, improved quality of life, and greater care satisfaction among patients with chronic conditions.^22–25^

### Recruitment

We recruited ACHD patients and clinicians from within and outside the University of California, San Francisco (UCSF) to gather multi-regional perspectives. *Patients* were adults (≥18 years) who spoke English or Spanish and could provide informed consent. Those with a developmental disability were not eligible. Initially, patients were identified through a retrospective chart review at UCSF. CHD diagnosis was documented for each participant and categorized as complex, moderate, or simple according to the published ACHD guidelines.^26^ Patients with isolated simple lesions, such as atrial septal defects or ventricular septal defects, were excluded because the guidelines regarding their frequent follow-up with ACHD specialists are less robust.^26^ We purposefully sampled^27^ patients who lacked an established relationship with an ACHD cardiologist or who had not seen one for at least three years during adulthood.^6,26^ We made deliberate efforts to include participants who were difficult to reach, as these individuals may face similar barriers to accessing specialized ACHD care. After thematic saturation^28^ was reached among UCSF patients, we recruited patients from outside UCSF through convenience and snowball sampling to ensure representation from all regions of the US (Northeast, Southeast, Midwest, Southwest, and West). We provided contact information for the study and/or our recruitment flyer to the CHD physicians in our network across the US, and to the members of the advocacy organization, Adult Congenital Heart Association (ACHA), to refer patients with CHD for interviews. While CHD diagnoses for UCSF patients were determined via chart review, non-UCSF patients provided their diagnoses verbally via self-report. *Clinicians* were CHD cardiologists or other CHD clinic staff members (social workers, nursing staff, patient care coordinators, etc.). We identified and recruited clinicians within UCSF based on their involvement with pediatric or adult CHD clinical care. Clinicians nationwide were identified through collaboration with the ACHA and snowball sampling within professional and community networks to represent all regions of the US.

### Data Collection

Separate patient and clinician interview guides were developed, incorporating the COM-B model to explore the barriers and facilitators to the uptake of specialized ACHD care. Semi-structured interviews were conducted over Zoom, each lasting 45-60 minutes. At the end of the interview, participants were given a recap of the major themes recorded to provide a member check.^29^ Most interviews were conducted in English by a single interviewer (KM). Four interviews with Spanish-speaking participants were conducted by a second interviewer (KB). Interviews were audio-recorded, professionally transcribed, and translated (when applicable). Patients completed a demographic survey (age, gender identity, race/ethnicity, highest level of education, estimated household income). They answered a single closed-ended question about the presence or absence of a prior three-year (or greater) gap in seeing a cardiologist.^26^

### Data Analysis

The coding team consisted of AA (ACHD cardiologist with formal qualitative research training), KM (medical student), and research assistants (JM, KB, KS, PA, AQ). All team members completed structured training in qualitative coding principles, including group practice coding of pilot transcripts to establish shared understanding of code definitions and application. Five coders (AA, KM, JM, KB, KS) collectively reviewed the first six (3 patient and 3 clinician) interviews using inductive coding to identify coding categories and subcategories of determinants of specialized ACHD care. We then conducted deductive coding using the developed list of thematic categories and subcategories. Each transcript was coded separately in Microsoft Word by two members of the research team (AA, KM, JM, KB, KS, PA, AQ). The two independently coded versions were compared in iterative consensus meetings to identify and resolve discrepancies, with guidance from more established qualitative researchers (MO, DD).

The consensus process generated a single, finalized coded transcript for each interview. Coded excerpts were organized in a Microsoft Excel spreadsheet, creating a matrix of themes and subthemes across participants. The matrix was analyzed using Rapid Qualitative Analysis,^30^ an action-oriented method for analyzing qualitative data, to efficiently produce results for real-world interventions. Using the matrix, cross-participant comparisons identified patterns in determinants of ACHD specialized care and behavioral factors. Summarizing matrix findings occurred via iterative group meetings of five team members (AA, KM, KB, JM, PA).

This research was approved by the UCSF Institutional Review Board (#22-36667). All participants completed verbal informed consent. The principal investigator, Dr. Agarwal, has full access to all study data and takes responsibility for its integrity and the data analysis. Participants initially received a $25 gift card per interview which was increased to $50 to enhance recruitment.

## III. Results Participants

As described in Table 1, we interviewed 37 patients between 20 and 56 years old; most were women (57%), monolingual English speakers (70%), and people of color (62%). Approximately 30% of patient participants required three or more outreach attempts to complete the interview, underscoring the difficulty of engaging this population and the importance of persistent, proactive recruitment strategies in ACHD. Clinicians (n=17) who participated in interviews were mostly ACHD cardiologists (41%), or nurse practitioners (24%), and 52% were from UCSF.

**Table 1:** Participant Characteristics Patient Characteristics (N=37)

| Table 1: Participant Characteristics |  |
| --- | --- |
| Patient Characteristics (N=37) |  |
| Age (years) |  |
| Median Age (25<sup>th</sup> percentile, 75<sup>th</sup> percentile)□ | 32 (25,38)□ |
| Age Range□ | 20-56 |
| Gender Identity, n (%) |  |
| Woman | 21 (57%) |
| Man | 15 (40%) |
| Non-Binary | 1 (3%) |
| **Race/Ethnicity\*, n (%)** |  |
| Alaska Native or Native American | 1 (3%) |
| Asian | 7 (19%) |
| Black/African American | 5 (14%) |
| Hawaiian or Pacific Islander | 1 (3%) |
| Hispanic/Latino | 9 (24%) |
| White | 13 (35%) |
| Other** | 2 (5%) |
| **Language, n (%)** |  |
| Monolingual English | 26 (70%) |
| Monolingual Spanish | 4 (11%) |
| Bilingual (English + another language)*** | 7 (19%) |
| **Geographic Location, n (%)** |  |
| Midwest | 2 (5%) |
| Northeast | 2 (5%) |
| Southeast | 3 (8%) |
| Southwest | 2 (5%) |
| West (not including UCSF patients) | 1 (3%) |
| West (UCSF patients) | 27 (73%) |
| **Highest Level of Education, n (%)** |  |
| Some high school, no diploma | 5 (14%) |
| High school graduate/GED/alternative | 5 (14%) |
| Some college, no degree | 5 (14%) |
| Bachelor's degree | 13 (35%) |
| Graduate/professional degree | 8 (22%) |
| Decline to answer | 1 (3%) |
| **Estimated Annual Household Income, n (%)** |  |
| Under \$20,000 | 4 (11%) |
| \$20,001 - \$40,000 | 4 (11%) |
| \$40,001 - \$60,000 | 3 (8%) |
| \$60,001 - \$80,000 | 3 (8%) |
| \$80,001 - \$100,000 | 2 (5%) |
| \$100,001 + | 10 (27%) |
| Decline to answer | 11 (30%) |
| **Prior Gaps in Care, n (%)** |  |
| Yes (3 years or more) | 13 (35%) |
| No | 24 (65%) |
| **CHD Complexity, n (%)** |  |
| Complex or Moderate | 18 (49%) |
| Simple | 13 (35%) |
| Unknown | 6 (16%) |
| **Clinician Characteristics (N=17)** |  |

| **Role/Specialty, n (%)** |  |
| --- | --- |
| Pediatric Cardiologist | 3 (18%) |
| Adult CHD Cardiologist | 7 (41%) |
| Nurse Practitioner | 4 (24%) |
| Allied Health Professional | 3 (18%) |
| **Geographic Location, n (%)** |  |
| Midwest | 2 (12%) |
| Northeast | 1 (6%) |
| Southeast | 0 (0%) |
| Southwest | 1 (6%) |
| West (not including UCSF clinicians) | 4 (24%) |
| West (UCSF clinicians) | 9 (52%) |
| Abbreviations: GED: General Educational Development (high school equivalency test); CHD: Congenital Heart Disease<br>*Multiracial individuals were counted for each racial/ethnic identity that they selected<br>**Includes those who described their race/ethnicity in a way not listed in the survey or declined to answer<br>***As determined by individuals who mentioned being bilingual during their interview. Not explicitly collected via survey data. |  |

### Barriers and Facilitators to ACHD Specialized Care: Intervention targets

Through iterative group analysis of coded transcripts, barriers and facilitators to specialized ACHD care clustered into six thematic categories. These categories emerged inductively from participants’ own language, experiences, and priorities. The six categories are described in the sections below and illustrated with representative participant quotes in **Table S1**.

### Healthcare System and Institutional Structure

This category describes broader structural factors that shape patients’ ability to access and remain engaged with specialized care. These include insurance, geographic location, scarcity of ACHD providers, and systemic policy (e.g. poor reimbursement models). Clinicians primarily identified this category as a high-priority intervention target, recognizing that structural barriers constrain engagement before individual motivation or knowledge can take effect. Patients described parallel challenges, including navigating insurance coverage and healthcare costs, adapting to changing policies, and retrieving childhood medical records essential to adult care. A distinct structural barrier emerged at the interface of pediatric and adult care systems: pediatric programs make repeated, persistent attempts to keep patients engaged, whereas adult care models more commonly close referrals after a few unanswered contacts, leaving recently transitioned patients at risk of falling through the cracks. Proposed interventions ranged from near-term, implementable solutions, including telehealth expansion and CHD-specific electronic health record alerts to identify and re-engage patients with care gaps, to longer-horizon systemic changes such as satellite clinic development, reimbursement reform for patient education visits, and ACHD workforce expansion.

> *“We cover about five different states and we’re the only ACHD program in our entire health system. We service a largely rural community and have patients who travel all over. It can be difficult when they live in a rural location — they don’t have the means or the understanding that they need to see us regularly.”* — Clinician ID #O_107

### Knowledge and Education

This category captures how patients and clinicians learn about CHD, understand the necessity of lifelong specialized follow-up, and acquire the practical skills to communicate effectively and navigate adult healthcare. Both patients and clinicians emphasized the need for centralized, regularly updated, and accessible resources that patients could engage with outside the clinic visit. Clinician-identified intervention targets included providing CHD-related education earlier in medical training, clinician education on navigating difficult conversations, patient education through innovative approaches (e.g., illustrations, videos, and 3D tools), and group educational sessions. Patients prioritized improved patient-physician communication regarding the transparency of their condition’s chronicity and CHD severity, individualized follow-up schedules, logistical guidance for day-to-day management of their CHD, and structured support at the point of transition, including both formal transition education and a deliberate provider handoff.

> *“I would like [information about my heart presented] verbally and in writing […] because sometimes, you know, we have a lot on our minds […] and then we forget, right? But if it’s written down, I could just read it again to remind myself and put it into practice”* – Patient ID #S_004

### Personal Development

This category captures shifts in a patient’s investment in their care based on developmental maturation, evolving motivation, and the gradual acquisition of self-advocacy skills over time. Both patients and clinicians noted that engagement is not static; it changes as patients grow older and encounter new life circumstances. Internal motivation and personal curiosity about one’s condition emerged as key facilitators, while developmental stage itself, particularly adolescence and young adulthood, was identified as a consistent barrier to engagement across participants.

This underscores the importance of developmentally tailored transition interventions that build self-advocacy skills progressively rather than transferring care responsibility abruptly at a fixed age.

> *“I think we just had a personal interest, to be honest — having a clinical interest in looking at results. When I have questions, I’m not asking ’Is everything okay?’ I would ask legit questions, like data and numbers. And I think it’s just a personal interest.”* — Patient ID #O_004

### Sources of Support

This category highlights the role of family, clinicians, and the broader CHD community in shaping patients’ engagement with their care. Family involvement emerged as a double-edged determinant: dependable support helped patients develop independent care management skills, while excessive caregiver involvement during childhood and adolescence could leave patients without the knowledge or confidence to navigate healthcare on their own. The sudden loss of a primary caregiver produced markedly different outcomes depending on the foundation of skills built prior to that loss. For some patients, it created significant barriers, and for others it catalyzed a rapid transition to independent care. Patient-clinician relationships were consistently described as powerful drivers of re-engagement. Peer and community support, such as formal mentorship programs, advocacy organizations, and faith communities, provided practical guidance and a sense of belonging that clinical encounters alone could not replicate.

> *“I mean my parents are still around, and they still kind of support me, but it’s my decision. It’s my actions to continue that kind of care. And because of how they were since Day 1, I continue that and make sure I’m asking the right questions, going to the right people.”* – Patient ID #O_006

### Identity and personal resources

This category encompasses the personal, cultural, and material circumstances that shape whether patients can access the healthcare system and feel comfortable and respected within it. Race, ethnicity, gender, language, socioeconomic status, comorbid conditions, and logistical barriers (including transportation, childcare, housing instability, and time off work) functioned as either barriers or facilitators depending on context. Clinicians identified logistical barriers as frequently unrecognized determinants of non-attendance requiring proactive screening and navigation support. Comorbid physical and mental health conditions further complicated engagement in ways that health systems are currently not consistently equipped to address.

A novel finding emerged regarding language in the context of non-English-speaking households. While language barriers in the clinical encounter itself remain important intervention targets, participants described how growing up in a household where parents had limited English proficiency sometimes paradoxically facilitated early patient engagement. In these households, children assumed responsibility for communicating with clinicians and scheduling appointments at a young age, thereby developing health navigation skills earlier than their peers.

> *“…because I grew up in an immigrant family and my parents don’t speak a lot of English, I had to have more understanding because I had [to] talk to the doctors who did not speak my language at home…Appointments, I started making early on. It used to be you had to call, so then I did the calling even at a young age, like before 18.”* – Patient ID #O_008

### ACHD placement in one’s life

This category signifies how patients balance the time, attention, and emotional weight of their condition against competing life priorities, and the degree to which they have integrated their CHD into their sense of self. Patients described a spectrum of relationships with their condition, from active avoidance and denial to acceptance and integration, and recognized that their position on this spectrum shifted across their lifespan in response to health status changes and life transitions. Anxiety and psychological burden drove avoidance of care for some patients, while family and personal legacy served as powerful motivators for others. Competing priorities such as career and financial pressures led some patients to deprioritize follow-up during periods of perceived stability.

> *“It can be really easy to wanna get sucked up in regular life. But it has to be the core of my life. I tell my friends I’m kind of at the mercy of my heart thing.”* — Patient ID #O_003

### Interdependence of Categories

The six categories were not mutually exclusive. Barriers or facilitators in one category frequently intersected with and compounded those in another. Structural factors requiring broader systemic solutions influenced factors typically considered within a patient’s control, such as their attitude toward follow-up care. The following account from a single patient illustrates how barriers across categories can compound one another:

> *“Having these symptoms and not getting answers is just giving me a lot more anxiety… It is affecting my income and employment, because it’s affecting my ability to work…the insurance is something that’s really preventing me from receiving the care, because I feel the doctors are being discriminatory because of my insurance…The ACHD provider availability, I was never told that I should be followed…Knowledge, I barely have. The only knowledge I have of my heart condition is really what I’ve read online.”* – Patient ID #U_016

Given this interdependence, interventions identified within three of the six thematic categories emerged as the most practical, readily actionable, and potentially high-impact solutions to address barriers present across some other categories: healthcare system and institutional structure, knowledge and education, and sources of support. Interventions in these three categories, such as reimbursement for educational visits, centralized and updated patient resources, and structured peer support programs, can be designed and implemented using relatively accessible approaches, while their effects are likely to extend across the other three categories (personal development, identity and personal resources, ACHD’s placement in one’s life), given the interconnected nature of the categories.

### Development of the Theory-Based Framework for Improving ACHD Care

We mapped the six categories onto the COM-B and TDF behavioral frameworks to clarify not just *what* to target, but *how* and *why* interventions in those domains could produce meaningful change in patient behavior (**Table 2**). For example, interventions targeting knowledge and education operate primarily through the TDF domains of Knowledge, Cognitive and Interpersonal Skills, Memory and Decision Processes, and Behavioral Regulation - all of which map to the Capability component of COM-B. This means that a knowledge-focused intervention would need to do more than transfer information; it would also need to build skills, support retention, and help patients develop self-regulatory habits around care engagement. Similarly, interventions targeting sources of support operate through Social Influences, Reinforcement, and Emotions - mapping to Opportunity and Motivation domains - meaning they work by changing the relational and emotional context of care rather than by building individual knowledge or skills.

**Table 2:** Mapping of ACHD Engagement Categories to the Theoretical Domains Framework (TDF)

|  |  |  |
| --- | --- | --- |
| **Category** | **What This Category Captures Behaviorally** | **TDF Domain(s) (*definition*)** |
| Healthcare System and Institutional Structure | Structural conditions that expand or constrain patients' physical access to specialized ACHD care, independent of individual motivation or knowledge | - Environmental Context and Resources (*any circumstance of a person's situation or environment that discourages or encourages the development of skills, independence, or adaptive behavior*) |
| Knowledge and Education | Patients' and clinicians' capacity to understand, remember, and act on information about CHD diagnosis, management, and follow-up | - Knowledge (*awareness of the existence of something*);<br>- Cognitive and Interpersonal Skills (*an ability or proficiency acquired through practice*);<br>- Memory, Attention and Decision Processes (*the ability to retain information, focus selectively on aspects of the environment, and choose between alternatives*);<br>- Behavioral Regulation (*anything aimed at managing or changing objectively observed actions*) |
| Personal Development | The internal drive, skills, and developmental readiness that enable patients to take increasing ownership of their care over time | - Beliefs about Capabilities (*acceptance of the truth or validity of one's own ability to perform a behavior*);<br>- Intentions (*a conscious decision to perform a behavior or resolve to act in a certain way*);<br>- Goals (*mental representations of outcomes or end states that an individual wants to achieve*);<br>- Optimism (*confidence that things will happen for the best or that desired goals will be attained*);<br>- Social/Professional Role and Identity (*a coherent set of behaviors and displayed personal qualities of an individual in a social or work setting*) |
| Sources of Support | The interpersonal relationships and community connections that reinforce or undermine a patient's sustained engagement with specialized care | - Social Influences (*interpersonal processes that can cause individuals to change their thoughts, feelings, or behaviors*);<br>- Reinforcement (*increasing the probability of a response by arranging a dependent relationship between the response and a given stimulus*);<br>- Emotions (*complex reaction patterns by which an individual attempts to deal with a personally significant matter or event*);<br>- Social/Professional Role and Identity (*defined above*) |
| Identity and Personal Resources | The personal, cultural, and material circumstances that shape whether patients can access and feel comfortable and respected | - Environmental Context and Resources (*defined above*);<br>- Social/Professional Role and Identity (*defined above*); |
|  | within the healthcare system | - Emotions (*defined above*);<br>- Beliefs about Capabilities (*defined above*) |
| ACHD's Placement in One's Life | The degree to which patients integrate their CHD into their sense of self and daily priorities, influencing their sustained motivation to engage in care | - Beliefs about Consequences (*acceptance of the truth or validity about outcomes of a behavior in a given situation*);<br>- Optimism (*defined above*);<br>- Goals (*defined above*);<br>- Emotions (*defined above*);<br>- Behavioral Regulation (*defined above*) |
| Definitions for TDF domains are adapted from Atkins et al., 2017<sup>18</sup>. The relationship between TDF domains and COM-B components is illustrated in Figure 1. Where a TDF domain appears in more than one category, its definition is provided at first mention and noted as "defined above" thereafter. The three categories with the greatest intervention opportunity: Healthcare System and Institutional Structure, Knowledge and Education, and Sources of Support - are highlighted in blue. |  |  |

Synthesizing across these frameworks, using patient activation and engagement as the central behavioral target, we developed the ACHD Care Wheel framework to support the design and evaluation of future interventions (**Figure 2**). The wheel is organized into four concentric layers, read from the outside in. The outermost layer represents all six categories of determinants of specialized ACHD care. These act on the second layer - TDF domains grouped into four conceptual clusters: Capacity to Act, Sense of Self, Social and Emotional Drivers, and Context and Consequences. Activation of these clusters shapes the third layer - the Capability, Opportunity, and Motivation - which together converge on the innermost layer of ACHD patient activation and engagement behavior. Using the chronic care model,^22^ we hypothesize that interventions supporting patient activation and engagement will improve ACHD specialized care, reduce emergent hospitalizations, increase patient satisfaction, and improve quality of life.^22–24^

**Figure 2:**
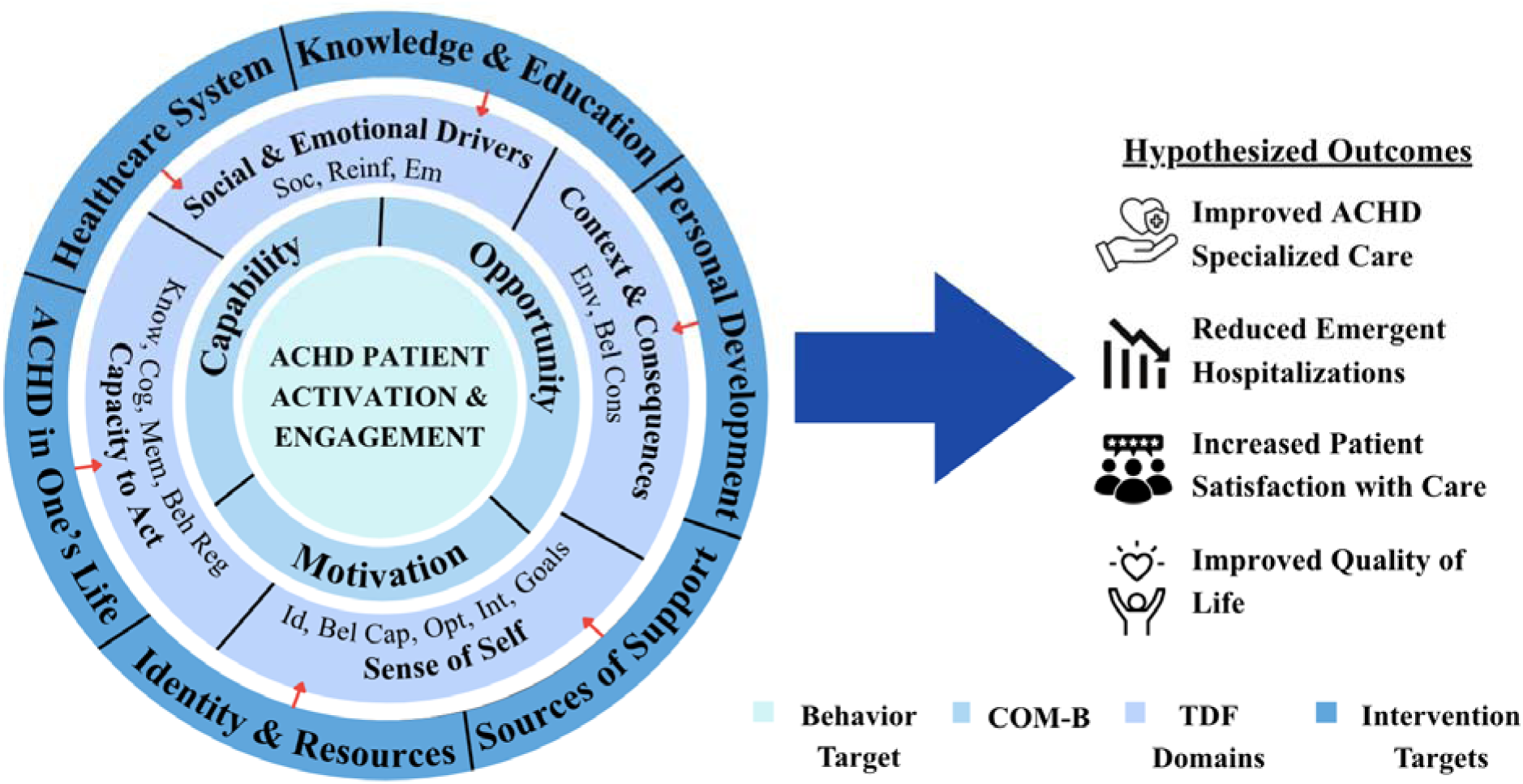
ACHD Care Wheel: A Stakeholder- and Theory-Grounded Framework for Improving Specialized ACHD Care. The outermost ring represents the six categories of determinants (listed using abbreviated labels) of specialized ACHD care identified from qualitative interviews with ACHD patients and clinicians. The middle ring represents relevant Theoretical Domains Framework (TDF) domains. TDF domains are grouped into four conceptual clusters: Capacity to Act; Sense of Self; Social and Emotional Drivers; and Context and Consequences. The inner ring represents the COM-B components of Capability, Opportunity, and Motivation. Together these converge on the central target behavior: ACHD patient activation and engagement which are hypothesized to improve specialized ACHD care and outcomes. The wheel’s layers are designed to rotate independently: any intervention target in the outer ring can align with any TDF cluster and COM-B component depending on the intervention’s design, rather than implying fixed mappings between adjacent segments. *Abbreviations: Soc, Social Influences; Reinf, Reinforcement; Em, Emotion; Env, Environmental Context and Resources; Bel Cons, Beliefs about Consequences; Id, Social/Professional Role and Identity; Bel Cap, Beliefs about Capabilities; Opt, Optimism; Int, Intentions; Goals, Goals; Know, Knowledge; Cog, Cognitive and Interpersonal Skills; Mem, Memory, Attention and Decision Processes*

## IV. Discussion

We describe an in-depth understanding of the patient and clinician perspectives on how ACHD specialized care could be improved. Six categories emerged as determinants for specialized care: healthcare system and institutional structure, knowledge and education, sources of support, personal development, identity and personal resources, and ACHD’s placement in one’s life.

From these findings we developed the novel ACHD Care Wheel, a stakeholder- and theory-driven framework for developing and evaluating interventions to improve specialized ACHD care.

### The ACHD Care Wheel as a Tool for Intervention Design and Evaluation

The ACHD Care Wheel can be applied in two directions. Prospectively, clinicians, health systems, or researchers designing a new intervention can enter the wheel at the outermost ring to select a target category. They can then trace inward to identify which TDF domains and COM-B components the intervention is intended to activate. Because the wheel’s layers rotate independently, an intervention entering at any outer category can connect to any TDF cluster and COM-B component. This clarifies not only what the intervention targets, but how and why it is expected to produce change. Consider a formal pediatric-to-adult transition program that builds patients’ knowledge of their condition and connects them with adult specialists. This maps onto the Knowledge and Education and Sources of Support categories. Tracing inward, the wheel makes explicit that such a program should activate the Capacity to Act cluster (Knowledge, Cognitive and Interpersonal Skills, Memory and Decision-Making skills), the Social and Emotional Drivers cluster (Social Influences), and Sense of Self cluster (Believe about Capabilities) engaging the Capability, Opportunity, and Motivation components, and evaluation should measure exactly that.

Retrospectively, the wheel helps explain why an existing intervention fell short. If a transition program were found ineffective, the wheel points to specific possibilities: perhaps it build knowledge without addressing patients’ retention or decision-making skills (Capability) or confidence to act on the knowledge (Motivation), or connected patients to a specialist who remained inaccessible due to insurance or geography (Opportunity). The wheel localizes the gap rather than treating the intervention as a uniform success or failure. Because the six categories are interdependent, an intervention designed around one category is likely to activate TDF clusters and COM-B components associated with others. Interventions centered on knowledge and education, such as reimbursement for educational visits or centralized patient resources, can also address barriers in personal development and ACHD’s placement in one’s life. Well-designed interventions in priority categories therefore need not address every category independently; their effects are likely to propagate across the framework and compound their overall impact.

### Prioritization of Intervention Targets

Three intervention targets stood out as the most practical, readily actionable, and potentially high impact: healthcare system and institutional structure, knowledge and education, and sources of support. Patients and clinicians differed, however, in both the priority and the proposed implementation of each. Clinicians focused on macro-level systemic interventions such as reimbursement reform, workforce expansion, and EHR-based alerts, whereas patients centered micro-level relational factors such as their connection with their provider and a sense of community belonging. These differences likely reflect a patient’s personal lived experience with CHD compared to a clinician’s experience treating many patients, and they suggest that effective interventions will need to operate at both levels simultaneously: system-level changes that remove structural barriers, and relationship- and community-centered approaches that address the personal determinants patients themselves prioritize.

The remaining categories (personal development, identity and resources, and ACHD’s placement in life) were viewed as more challenging and resource intensive to target. This reflects their intrinsic, personal nature, which complicates designing interventions for the diverse ACHD community, and a perceived inability to address structural inequities in resource allocation treatment based on race, ethnicity, culture, language, and comorbidities with smaller-scale interventions. Participants identified these factors as crucial to improve care, yet solutions of this scale may seem too distant to design and implement within current resource constraints.

### Novel Findings: Language and Caregiver Support as Complex Determinants

Two findings, related to language and caregiver support, extend current understandings of barriers to ACHD engagement. Prior research consistently frames language barriers as impediments to care, with non-English-speaking households more likely to miss appointments and disengage from clinicians.^31,32^ Our findings add nuance to this. While language barriers in the clinical encounter remain important intervention targets, participants described how growing up in a household where parents had limited English proficiency sometimes facilitated early patient engagement. Children in these households assumed responsibility for communicating with clinicians and scheduling appointments before age 18, developing health navigation and self-advocacy skills earlier than their peers. Language barriers thus operate differently depending on whether they are located in the clinical encounter or in the patient’s household, a distinction with practical implications for interventions design. Similarly, dependable caregiver support helped many patients develop self-management skills, but excessive support during childhood and adolescence left some unable to navigate healthcare independently in adulthood, a vulnerability most apparent during sudden support disruptions such as loss of a primary caregiver. Together, these findings suggest that the quality and developmental appropriateness of language navigation and caregiver involvement may matter as much as their presence or absence. Structured approaches that progressively transfer care responsibility to patients beginning in adolescence represents a distinct and underutilized intervention target.

### Alignment with and extension of prior literature

Participant-prioritized solutions, such as multidisciplinary formal transition programs, are similar to those previously described.^29–33^ What distinguishes our approach is the application of COM-B and TDF to explain *how* proposed interventions could improve outcomes in ACHD, and when an intervention falls short, to provide a principled basis for understanding why and how to improve rather than starting over. These frameworks were selected because they enable mechanism-based recommendations for intervention design targeted to the desired behavior.^17–19^ Theoretical grounding does not guarantee success, but it increases the likelihood that proposed solutions will work by targeting the behavioral determinants most likely to drive change and by drawing on evidence of what works, for whom, and under what circumstances.

### Limitations

Some ACHD stakeholder perspectives may not be captured in our sample. We reduced this likelihood by recruiting a large, diverse sample of both patients and clinicians across social and geographic domains. Although 73% of patient participants were recruited from UCSF, non-UCSF recruitment continued until thematic saturation and geographic representation across all five US regions were achieved. Findings should nonetheless be interpreted with awareness of the academic medical center context of the majority of our sample. While we used COM-B and TDF, other behavioral frameworks may be equally or better suited to capture the complexities of how patients interact with their health and healthcare, and future work can explore integrating them. Finally, a different interviewer for Spanish-speaking participants may have introduced bias, though this was mitigated through a shared interview guide and consensus-based coding. The benefit of including monolingual Spanish speakers was judged to outweigh this risk.

## V. Conclusion

We present a stakeholder- and theory-grounded framework for improving specialized ACHD care, developed from the perspectives of diverse patients and clinicians across the United States. Three intervention categories emerged as the most actionable and potentially high-impact targets: healthcare system and institutional structure, knowledge and education, and sources of support. The ACHD Care Wheel provides a structured, transparent scaffold for designing and evaluating interventions; clarifying what to target and how and why change is expected. Given the interdependence among categories, well-designed interventions are likely to produce effects across multiple domains simultaneously, amplifying their overall impact. Future work should apply the ACHD Care Wheel prospectively, beginning with transition programs, where the need is most acute and the framework’s multilevel logic is most directly applicable, with the goal of reducing the preventable morbidity and mortality that results from gaps in specialized ACHD care.

## Data Availability

All data produced in the present study are available upon reasonable request to the authors

## Non-Standard Abbreviations and Acronyms

ACHA: Adult Congenital Heart Association
ACHD: Adult Congenital Heart Disease
CHD: Congenital Heart Disease
COM-B: Capability, Opportunity, Motivation–Behavior
TDF: Theoretical Domains Framework
UCSF: University of California, San Francisco

