## Supplemental Table 1 for "ACHD Care Wheel: Stakeholder- and Theory-Driven Framework to Improve Specialized Adult Congenital Heart Disease Care"

**SUPPLEMENTAL MATERIAL**

| **Table S1. Determinants of ACHD Patient Engagement in Specialized Care with Illustrative Quotes** | | |
| --- | --- | --- |
| **Category** | **Subcategory** | **Exemplar Quotes** |
| **Healthcare System and Institutional Structure** | Navigating Insurance and Healthcare Costs | “I think that was the part that made me more nervous than the actual transition to adult care was, more just the, ‘Can I afford this?’ I need to afford it. It’s my health. It’s important. I have to make it work. But it was more of, ‘How am I going to do this?’” - Patient ID #O_007 |
|  | Overcoming Geographic Distance and Accessing ACHD Providers | “Location and availability of ACHD providers. There is only a few hundred ACHD centers that are ACHA accredited and have more than one ACHD provider working…If you have to drive several hundred miles to get to an ACHD provider, that’s gonna be a huge barrier. It’s time, it’s cost, it’s energy”- Clinician ID #O_102 |
|  | Addressing Systemic and Policy-Level Barriers to ACHD Care | “it seems to me as if it’s always changing from privacy rules and how information is shared across other physicians.” - Patient ID #U_007 |
|  |  | “I would have had a third open-heart surgery like four years ago. And COVID happened so it delayed it”- Patient ID #U_009 |
|  |  | “The learning curve is steep. And then at the end of all of it, you don’t get paid any more than a general cardiologist…And you’re non-invasive. You’re not a proceduralist who can make money for the institution and bill higher RVUs for procedures.”- Clinician ID #O_102 |
|  | Retrieving Past Medical Records | “it’s very important that we have our records accessible.  Like right now – I had surgery at [a] children’s hospital 47 years ago. You will never get those records, and there are pieces of those records that they actually need now to answer questions that they have about my valve, but they’ll never get those answers.” - Patient ID #O_001 |
|  | Bridging Pediatric and Adult Care Models | “The other thing with [the] adult world that we’ve seen is someone will call the family or patient three times, and if the patient doesn’t respond, they close the referral, which, we can call our patients 20 times just to get them into a clinic visit. We don’t close the referral. We don’t close their access to care, and I think that the adult model is very different than the pediatric model. We do a lot of handholding, but it really is support.” - Clinician ID #U_104 |
| **Knowledge and Education** | Understanding One's CHD Diagnosis and Care Needs | “I’m sometimes not able to really explain or know the name of my heart condition. I don’t know the extreme of how complicated it is. As far as the medicine…sometimes I forget, and sometimes I need reminders as to why I have to continue the medicine and the purpose of it.”- Patient ID #U_017 |
|  | Receiving Structured Transition | “I did a Zoom thing with her, and…she helped me transition. She was like, ‘This is what we’re doing, and here’s your new doctor.’ …She’s like, “Oh, what’s your job? Or what are you studying to be? And make sure the job you have has the good health insurance.’…it was basically like an introduction Zoom meeting of ‘Now, you’re an adult, and she’s here to help me…transition into the adult sector.’” - Patient ID #U_008 |
|  |  | “You have them meet with the team at one appointment…Something where you can introduce the provider – like, the patient’s current provider can introduce the other provider they’ll be seeing”- Clinician ID #U_109 |
| **Personal Development** | Developing Internal Motivation and Self-Advocacy | “it takes a unique patient, it takes a unique attitude, and you have to be really relentless to want to navigate the healthcare system. You can’t do it passively. You have to be very active in doing it.”- Clinician ID #O_103 |
|  |  | “Reading material online is basically the only way I could really help advocate for myself. It’s the only way I’ve been able to advocate for myself with my doctors” - Patient ID #U_016 |
|  | Embracing Responsibility for Care as One Matures | “I guess I grew up. And I finally realized what I have to do in life and have to grow up now. I have a situation. And I have to really deal with it and be a man about it and just grow up” - Patient ID #U_010 |
|  |  | "Patient engagement is much more difficult in most populations. Then you add onto it the fact that they're teenagers or young adults - I think they don't really want to engage." - Clinician ID #O_103 |
| **Sources of Support** | Navigating Family Involvement in Care | “my mom took me, and she helped me fill out the papers and all that.” – Patient ID #S_004 |
|  |  | “I see a lot of times where our parents – the parents of kids with ACHD – are almost too involved in care in adolescence…Many times, questions are asked of the patients and the parents answers. And then, unfortunately they do this all the time, and the patient ends up growing into adulthood and not really having a full understanding of what’s going on with their heart condition.”- Clinician ID #O_107 |
|  | Drawing on Family as a Motivator for Engagement | “My kids inspire me. I want to be with them when I’m old.” - Patient ID #O_010 |
|  | Connecting with CHD Peer, Community, and Spiritual Support | “I looked into that and saw that they had this peer mentor program. And I thought, well, that might be really good to just talk with someone who’s gone through something similar at least, surgery as an adult. So, I reached out and got paired with someone.”- Patient ID #O_006 |
|  |  | “And so, with prayer and stuff, just with, I would say honestly God is the one that healed me”- Patient ID #U_020 |
|  | Building a Trusting Patient-Clinician Relationship | “initially I didn’t wanna go to the doctors. And now, I feel that I have a connection with the new doctors that I know” - Patient ID #O_010 |
| **Identity and Personal Resources** | Navigating Identity-Based Barriers and Facilitators in Clinical Encounters | “I actually like the diversity at the front desk and the staff. It makes me feel comfortable… It just feels good to be welcomed by a variety of different people”- Patient ID #U_017 |
|  |  | “It was really important for me to have a woman provider, because I have had male providers, and I just did not have good experiences with them listening to me.” - Patient ID #U_018 |
|  | Managing Comorbid Physical and Mental Health Conditions | “I have PTSD and OCD from an unrelated thing. And I think that I just struggle with anxiety and tasks that give me anxiety. Especially the whole thing where I was getting sicker and sicker and dealing with insurance.” - Patient ID #U_021 |
|  |  | “I do self-medicate with methamphetamines, but that is because I have not been able to get my ADHD medication. And all my doctors are telling me what I’m using is what’s causing all my symptoms, and they’re not reassuring me of anything.” - Patient ID #U_016 |
|  | Overcoming Socioeconomic Barriers to Care | “The ones who express an understanding, what their heart conditions are, at least the ones who I think higher percentage-wise, are gainfully employed. And even if they lose jobs, they're getting another job.” - Clinician ID #U_106 |
|  | Addressing Logistical Barriers to Attending Care | “for a while, I was homeless, and that was affecting my ability to maintain appointments.” - Patient ID #U_016 |
|  |  | “whether it’s finding out if they need rides to come and see us, finding out if they need childcare to come and see us, things that could help them find the time or ability to come and have better access to our center” - Clinician ID #O_104 |
| **ACHD’s Placement in One’s Life** | Reconciling Current Health Status with Need for Ongoing Care | “I just feel like maybe partially they feel like they’ve done a lot of procedures, they’re well for a certain amount of years, and then they just feel okay. And then they don’t come back…I think maybe a part of it is denial. Just not wanting to be sick anymore. Wanting to be normal.” - Clinician ID #U_108 |
|  | Managing Anxiety and Psychological Burden of CHD | “So, I have an anxiety disorder. Which as a kid I didn’t really have. So, it’s harder as an adult because I’m dealing with stress around it that I didn’t have as a kid.” - Patient ID #U_009 |
|  | Balancing CHD Care with Competing Life Priorities | “I think there was probably a time there when I was down in Southern California where I just let gaps occur just because I was starting a career, doing a lot of things.” - Patient ID #U_007 |
